# Repetitive Transcranial Magnetic Stimulation for Post-Stroke Rehabilitation: A Systematic Review and Meta-Analysis

**DOI:** 10.64898/2025.12.11.25342117

**Authors:** Forrest Lin, Roy H. Hamilton, Kelly L. Sloane

## Abstract

**Background:** Stroke is the leading cause of adult-onset disability, with impairments across motor, language, cognitive, swallowing, mood, and gait domains. Non-invasive brain stimulation techniques like repetitive transcranial magnetic stimulation (rTMS) have emerged as promising tools to augment rehabilitation therapies to improve post-stroke impairments. This systematic review and meta-analysis will evaluate the efficacy of rTMS for post-stroke recovery across multiple functional domains and identify moderators of treatment response.

**Methods:** We systematically searched five databases for randomized controlled trials (RCTs) published from February 16, 2004 to July 1, 2024. Eligible studies compared active rTMS with sham rTMS in stroke survivors and reported validated outcomes. Risk of bias was assessed using the Cochrane Collaboration tool. Random-effects meta-analyses were conducted for each outcome domain. Subgroup analyses examined timing of intervention (acute/subacute, early chronic, late chronic). Meta-regression evaluated continuous moderators, including stimulation intensity (% resting motor threshold), number of sessions, age, and sex.

**Results:** Fifty-two studies met inclusion criteria (2,472 participants, mean age 59.8 years; 35.1% female). rTMS was associated with significant improvements in motor function (UE-FMA, MD = 4.68, 95% CI [2.18, 6.54]), language (CCAT, MD = 0.62, 95% CI [0.22, 1.01]), cognition (MMSE, MD = 2.12, 95% CI [1.34, 2.92]), dysphagia (PAS, MD = –1.50, 95% CI [–2.40, –0.57]), and mood (HAMD-17, MD = –2.34, 95% CI [–4.38, –0.30]), but not gait. Subgroup analyses showed significantly larger treatment effects when rTMS was delivered within 3 months post-stroke, particularly for motor outcomes. Meta-regression indicated that stimulation intensity, number of sessions, and participant demographic distribution were not significant moderators.

**Conclusions:** rTMS improves post-stroke outcomes across multiple functional domains, with the strongest evidence for motor recovery in the acute/subacute phase. Standardization of protocols and larger trials in understudied domains are needed to maximize therapeutic outcomes.

## Introduction

Stroke is the leading cause of adult disability across the United States, with approximately 80% of stroke survivors experiencing impairments related activities of daily living.^1,2^ Post-stroke impairments in voluntary motor control are common and often persist beyond acute recovery, with over one half of survivors experiencing long-term motor deficits. These impairments contribute to learned nonuse, limit activities of daily living, and negatively affect return to work.^3^ Aphasia is a prevalent consequence of stroke, affecting approximately 30-40% of stroke survivors and impacting spoken language, auditory and reading comprehension, and writing.^4^ It is often associated with increased post-discharge healthcare costs and higher likelihood of recurrent stroke due to challenges in comprehending treatment instructions and communicating symptoms effectively.^5,6^ Though estimates vary based on assessment methods and timing, cognitive dysfunction remains a prevalent consequence of stroke, with nearly 75% of stroke survivors demonstrating cognitive impairment and one-third developing dementia within five years.^2,7^ Dysphagia, or impaired swallowing, affects more than half of survivors and is linked to heightened risks of pulmonary infections, malnutrition, and dehydration in individuals with persistent symptoms.^8,9^

The current standard-of-care for functional recovery involves rehabilitation with licensed therapy specialists targeting specific modalities. However, the effectiveness of this approach is often constrained by limited access, which is further limited by factors such as insurance coverage, affordability, and geographic availability of services.^10^ Other interventions aimed at improving post-stroke function have been investigated with limited success. Randomized controlled trials evaluating pharmacologic agents such as donepezil, escitalopram, and amantadine have reported only modest benefit.^11,12,13^ Similarly, virtual reality, computer-based training, and aerobic exercise have shown promising results in smaller studies, but large-scale randomized trials have yet to demonstrate consistent clinical benefit from these interventions.^14,15,16^ Thus, the lack of large-scale trials demonstrating efficacy has limited the implementation of methods to address post-stroke impairments.

Recovery of function after stroke is supported by neuroplastic processes that occur spontaneously in the immediate post-stroke period.^17,18^ Neuroplasticity, the brain’s innate ability to reorganize its structure and function in the face of physiologic damage, drives neural reorganization and synaptic changes. Non-invasive brain stimulation techniques, like transcranial magnetic stimulation (TMS), offer the potential to enhance spontaneous recovery by influencing neural network activity. TMS has emerged as a promising adjunct to rehabilitation by enhancing neuroplasticity and optimizing recovery outcomes. This review explores the potential of rTMS in post-stroke rehabilitation by performing a meta-analysis of available randomized controlled trials (RCTs). It evaluates the efficacy of rTMS across multiple recovery domains, including motor, cognitive, language, dysphagia, gait, and post-stroke depression, and synthesizes current evidence on its application to treatment outcomes. The review further aims to provide a comprehensive understanding of when rTMS is most effective in promoting recovery after stroke.

### Mechanism of Action of TMS

Transcranial magnetic stimulation (TMS) operates by transmitting an electric current through conductive wires within an insulated coil, generating a localized magnetic field that passes through the skull with minimal resistance, inducing a secondary electric field in the underlying cerebral cortex. The resulting current causes neuronal depolarization, exciting or inhibiting the cortex.^19^

Repetitive TMS (rTMS) refers to an approach that delivers a series of magnetic pulses to repetitively depolarize targeted neurons and thereby induce long-lasting changes in neuronal activity. The effects of rTMS depend on the frequency and pattern of stimulation. Non-patterned rTMS is commonly used to modulate brain activity by applying magnetic pulses at consistent, sustained frequencies. High-frequency non-patterned rTMS typically involves delivering magnetic pulses at frequencies between 10-20 Hz. This type of stimulation is associated with increased cortical excitability and facilitation of long-term potentiation (LTP), a mechanism linked to synaptic strengthening and functional recovery.^20,21^ Low-frequency rTMS involves applying stimulation at a slower rate, usually below 5 Hz. It is associated with a decrease in cortical excitability and long-term depression (LTD)-like effects.^22^

Theta burst stimulation (TBS) is a patterned variant of rTMS that delivers short bursts of high-frequency pulses (∼50 Hz) that are repeated at regular intervals (5 Hz). TBS mimics the brain’s endogenous theta rhythms known to drive synaptic plasticity, enabling it to more precisely engage LTP- or LTD-like mechanisms.^23^ TBS protocols can be divided into two variants: intermittent theta burst stimulation (iTBS) and continuous theta burst stimulation (cTBS). iTBS is administered by delivering bursts of 3 pulses at 50 Hz every 200 milliseconds, repeated for 190 seconds (600 pulses in total). iTBS promotes LTP-like plasticity, supporting synaptogenesis and functional recovery. In contrast, cTBS uses the same burst pattern (3 pulses at 50 Hz), but delivers the bursts continuously for ∼40 seconds without break (600 pulses total). cTBS produces LTD-like effects that suppress synaptic transmission and reduce excitability.^24^

Because rTMS can create a state of heightened plasticity in targeted networks, functional recovery has been shown to be significantly amplified when paired with a targeted training task.^25^ Task-dependent plasticity leverages the principle that synaptic modifications are most robust when neural stimulation coincides with behaviorally relevant activity, thereby further reinforcing functionally meaningful synaptic connections.^26^ This approach allows rTMS to be tailored to specific goals of rehabilitation. While stimulation alters the excitability of a target region, concurrent training tasks shapes how that plasticity is functionally expressed. Consequently, paired protocols not only strengthen the effects of neuromodulation but also increase the relevance and durability of therapeutic gains.

### rTMS in post-stroke rehabilitation

The effectiveness of rTMS is dependent on stimulation target, paired training task, and method of stimulation. The choice of stimulation target and training task is determined by the functional modality being addressed, such as motor, language, or cognitive function, with each domain relying on activation of distinct neural networks. The method of stimulation is determined by the desired neurophysiological outcome, such as assessing cortical excitability, modulating inhibition, or inducing long-term potentiation (LTP) or long-term depression (LTD).

Many approaches to rTMS in post-stroke rehabilitation, particularly those targeting motor paresis, aphasia, and spatial neglect, are grounded in the interhemispheric imbalance model. This model assumes that in a healthy brain, the two hemispheres maintain a dynamic balance through mutual inhibition. Because the brain is organized in distinct vascular territories, damage following a unilateral stroke is usually confined to one hemisphere, disrupting this balance. As such, the affected (ipsilesional) hemisphere exhibits reduced inhibitory output, which leads to excessive excitability in the unaffected (contralesional) hemisphere. In turn, the contralesional hemisphere exerts excessive inhibition back to the ipsilesional hemisphere, further hindering recovery.^27^ Neuromodulatory techniques like rTMS can aim to restore this balance by either increasing excitability in the ipsilesional hemisphere or dampening hyperexcitability of the contralesional side.^28^

To enhance motor recovery for example, excitatory stimulation is often applied to the ipsilesional primary motor cortex while patients engage in motor-focused therapies such as constraint-induced movement therapy.^29^ In post-stroke aphasia, where damage often occurs in language centers of the left hemisphere, rTMS may enhance language recovery by exciting activity in the left hemisphere and suppressing the overactive right hemisphere. When combined with speech therapy, this approach has been shown to enhance naming accuracy and communicative function.^30^ Similarly in patients with spatial neglect, typically resulting from right hemisphere stroke, rTMS can be applied to stimulate the ipsilesional (right) hemisphere and inhibit the contralesional (left) hemisphere. This rebalancing of activity has been shown to improve visuospatial attention and cognitive function.^31^ While the interhemispheric imbalance model provides a theoretical foundation for some rTMS protocols, it does not fully capture the complexity of post-stroke cortical reorganization. Evidence highlights that recovery mechanisms are highly variable and are influenced by lesion location and individual neuroanatomy.^32^ Therefore, this model should be interpreted as one of several frameworks guiding neuromodulation strategies in post-stroke rehabilitation.

Observational studies of post-stroke recovery indicate that the period of heightened neuroplasticity occurring in the first 6-12 weeks after stroke correlates with spontaneous restoration of function.^33^ It is generally accepted that rTMS therapy administered within the first few months after stroke offers the most favorable outcomes, as brain plasticity is most pronounced during the early recovery phase. However, there remains a limited body of research specifically examining how the timing of rTMS intervention influences treatment outcomes in post-stroke patients. In this systematic review, we summarize the randomized controlled trials of rTMS in individuals with post-stroke impairments and provide evidence for optimal intervention timing in this population.

## Methods

### Literature Review

A systematic review was conducted using PubMed, EMBASE, PsycInfo, Cochrane Central Register of Controlled Trials, and Web of Science. The search encompassed all available studies prior to September 1st, 2024. Key search terms included “rTMS,” “RCT,” and “stroke,” using the Boolean search strategy: *((“rTMS” OR “magnetic stimulation”) AND (“randomized controlled trial” OR “RCT”)) AND (“stroke”).* No language restrictions were applied. All studies were published between February 16^th^, 2004 to July 1^st^, 2024.

### Eligibility Criteria

Randomized controlled trials (RCTs) enrolling post-stroke patients were included. Trials comparing an rTMS intervention group to a sham stimulation control group were included. Studies were required to specify the time since stroke onset for their participants and focus on a primary outcome modality, such as motor function, speech, cognition, mood, dysphagia, or gait. An initial screening of abstracts was assessed for eligibility using Covidence based on the predefined criteria.

### Data extraction and outcome measures

Covidence software was used to manage all aspects of screening, review, and extraction (www.covidence.org). Articles identified from the literature search were imported into Covidence, and duplicates were automatically removed. Two independent reviewers (FL and KS) screened each article based on title and abstract and subsequently performed full-text review. Any conflicts during the screening phases were resolved by a third independent reviewer. For crossover trials, only data from the first period (pre-crossover) were considered to minimize carryover effects. Included studies were first categorized based on their primary domain of interest: motor function, speech, dysphagia, cognition, mood, gait, or other. Studies were further stratified by time since stroke: acute/subacute (<3 months), early chronic (3–12 months), and late chronic (>12 months). For each study, the most prominent outcome measure within its respective domain was extracted (reported in ≥ 60% of included studies).

The following measures were extracted:

- Motor function: Upper extremity Fugl-Meyer Assessment (UE-FMA)
- Dysphagia: Penetration Aspiration Scale (PAS)
- Language: Concise-Chinese Aphasia Test (CCAT)
- Cognition: Mini-Mental State Examination (MMSE)
- Gait: 10-Meter Walk Test (10MWT)
- Mood: 17-item Hamilton Depression Rating Scale (HAMD-17)

In addition to primary outcome scores, data were extracted on potential effect modifiers, including participant characteristics (age, sex, ischemic to hemorrhagic stroke ratio) and rTMS intervention parameters (TMS device, coil location, stimulation frequency in Hz, % resting motor threshold, number of sessions, and number of pulses per session).

### Risk of bias and quality assessment

Risk of bias was assessed by FL and KS independently using the Cochrane Collaboration tool for assessing the risk of bias across six main domains: sequence generation, allocation concealment, blinding, incomplete outcome data, selective outcome reporting, other bias.^34^ Studies were subsequently categorized based on their overall risk of bias. Studies with high risk of bias were excluded from analysis to minimize the influence of methodological flaws on pooled estimates and to enhance the internal validity of the meta-analysis.

### Meta-analysis

Meta-analyses were performed separately for each outcome modality (motor, language, cognition, gait, dysphagia, mood) using the metafor package in R.^35^ A random-effects model was applied to account for heterogeneity across studies, including differences in methodology, sample characteristics, and the relatively small number of studies within each outcome modality. This model assumes that observed treatment effects may vary not only due to true differences between studies but also due to sampling variability or chance. For each modality, the primary effect size was defined as the most commonly reported outcome measure specific to that domain. Effect sizes were calculated as the mean difference in scores between active and sham stimulation groups from baseline to post-intervention.

Statistical significance was evaluated using 95% confidence intervals (CIs) around the pooled effect size. Significance was inferred when the CI did not cross zero (p < 0.05). All included studies reported sufficient descriptive statistics (group means, standard deviations, and sample sizes), enabling direct calculation of standardized effect sizes. Between-study heterogeneity was assessed using Cochran’s Q statistic and the I² index. The Q statistic tests whether true differences in effect sizes (heterogeneity) exceeds what would be expected by random chance alone (homogeneity). I² quantifies the proportion of total variation across studies attributable to heterogeneity rather than sampling error. I² was reported because it provides a descriptive estimate of heterogeneity that is less dependent on the number of included studies than Cochran’s Q.

For outcome modalities represented by fewer than five studies, subgroup analyses were not conducted because such small samples violate statistical assumptions for reliable subgroup testing. Tests of interaction and heterogeneity are underpowered and prone to unstable estimates when study numbers are very limited.^36^ Subgroup analyses were conducted for categorical variables reported across studies to explore potential factors influencing treatment effects. The Q-statistic was used to compare treatment effects to time since stroke (acute/subacute, early chronic, late chronic). Meta-regression, a subgroup method that assesses moderator effects using regression-based techniques, was used to examine characteristics of continuous variables. These variables included stimulation intensity, number of treatment sessions and participant demographics (age and sex). Stimulation intensity was reported in each study as a percentage of the resting motor threshold (%RMT), ensuring that dose of stimulation is standardized relative to participant’s cortical excitability. Meta-regression was not performed for outcome modalities with five or fewer studies, as such analyses were underpowered due to limited degrees of freedom. Publication bias was assessed with Egger’s regression, which requires a minimum of 10 studies to provide stable estimates.^35^ For outcome domains with a limited number of studies, Begg’s rank correlation test was used to assess the risk of publication bias, as it is less influenced by small sample sizes. Institutional Review Board approval was not obtained because this meta-analysis was performed on previously conducted studies where data are published and publicly available.

## Results

The systematic search yielded 1640 publications. After removing duplicates and screening titles and abstracts, 242 full text studies were assessed for eligibility. After selection of articles following the flowchart described in **Figure 1**, the remaining 52 randomized clinical trials were included in the systematic review. In these studies, 2,472 participants were included – mean age of 59.8 years, 35.1% females. The main reasons for exclusion of full text studies were due to lack of sham protocol and failure to report the most common outcome measure. A risk of bias table is provided in **Supplementary Table 1** to evaluate the quality of data across the included studies. In total, there were 30 studies that included participants within 3 months since stroke, 12 studies that included participants within 3-12 months since stroke, and 10 studies that included participants more than 12 months since stroke.

**Figure 1.**
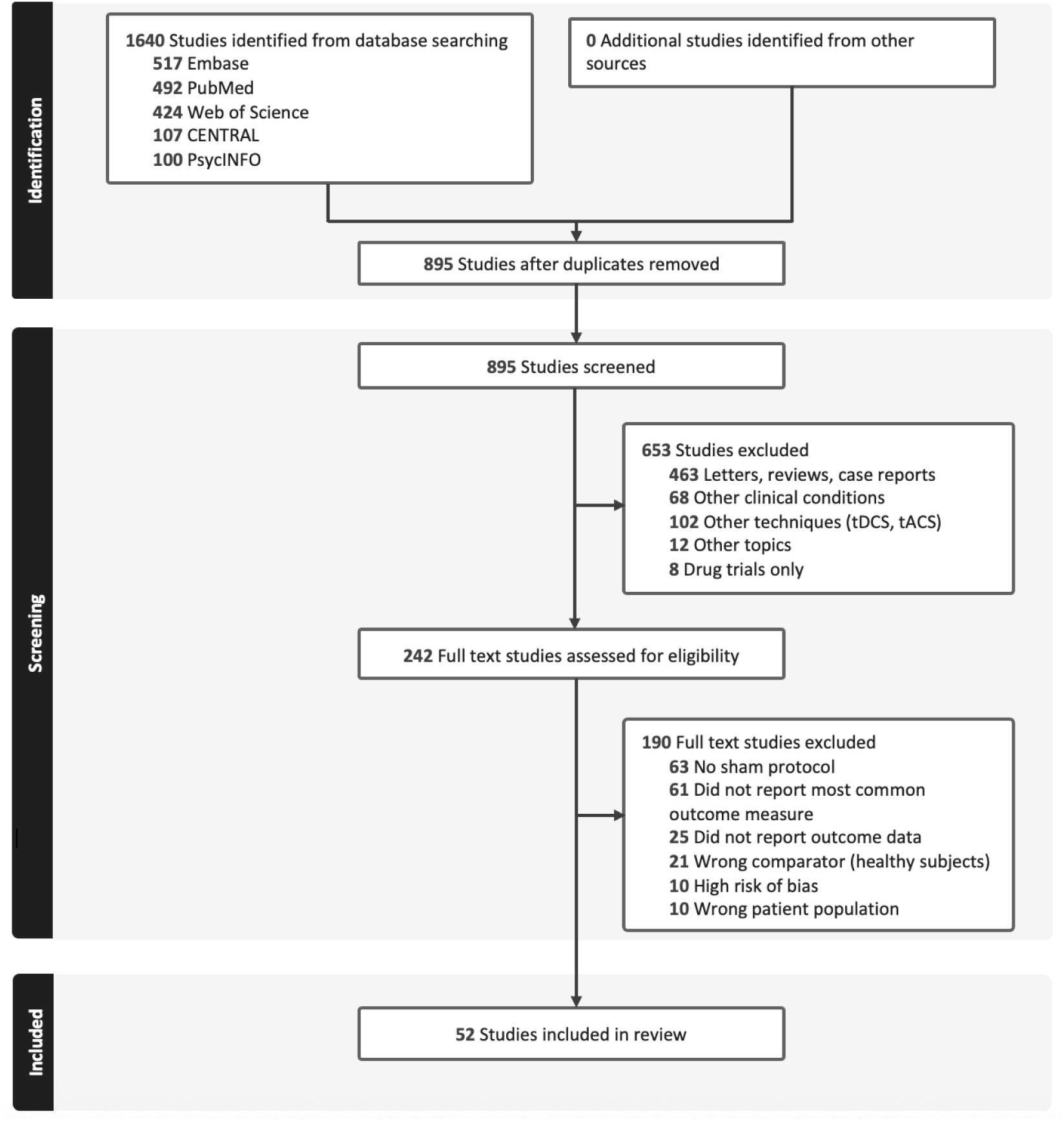
**PRISMA of flow diagram.**

### Heterogeneity across rTMS protocols by outcome modality

Across the observed outcome domains, substantial heterogeneity was observed in rTMS stimulation protocols, with variation in stimulation site, frequency, intensity, and number of sessions. A comprehensive summary of TMS parameters for each study, including stimulation site, frequency, number of pulses, percent motor threshold, and number of sessions, is provided in **Table 1**.

**Table 1.** Summary of study characteristics.

| ID | Study | Outcome Modality | Sample Size | Stimulation Site | Frequency (Hz) | # of Pulses | % Motor Threshold | Sessions |
| --- | --- | --- | --- | --- | --- | --- | --- | --- |
| [37] | Abdelkader et al., 2024 | Motor | 55 | Contralesional M1 | 5 | 1000 | 60 | 5 |
| [38] | Barros et al., 2014 | Motor | 20 | Contralesional M1 | 1 | 1500 | 120 | 10 |
| [39] | Blesneag et al., 2015 | Motor | 16 | Contralesional M1 | 20 | 1200 | 120 | 20 |
| [40] | Chang et al., 2010 | Motor | 28 | Ipsilesional M1 | 10 | 1000 | 90 | 10 |
| [41] | Chen et al., 2022 | Motor | 45 | Contralesional M1 | 1 | 1200 | 90 | 10 |
| [42] | Chen et al., 2021 | Motor | 23 | Ipsilesional M1 | iTBS (5 Hz bursts/50 Hz pulses) | 600 | 80 | 15 |
| [43] | Chen et al., 2019 | Motor | 22 | Ipsilesional M1 | iTBS (5 Hz bursts/50 Hz pulses) | 600 | 80 | 10 |
| [44] | Chieffo et al., 2014 | Gait | 9 | Bilateral lower limb M1 | 20 | 1500 | 90 | 11 |
| [45] | Chieffo et al., 2021 | Gait | 12 | Bilateral lower extremity M1 | 20 | 1500 | 90 | 11 |
| [46] | Chou et al., 2021 | Speech | 85 | left PTr | iTBS (5 Hz bursts/50 Hz pulses) | 600 | 80 | 10 |
| [47] | Duan et al., 2023 | Mood | 71 | Left DLPFC | 10 | 1400 | 80 | 20 |
| [48] | Edwards et al., 2023 | Motor | 60 | Contralesional M1 | 1 | 900 | 110 | 18 |
| [49] | Gottlieb et al., 2021 | Motor | 28 | Contralesional M1 | 1 | 1200 | 100 | 10 |
| [50] | Gu et al., 2017 | Mood | 24 | Left DLPFC | 10 | 1000 | 110 | 10 |
| [51] | Guan et al., 2017 | Motor | 42 | Ipsilesional M1 | 5 | 1000 | 120 | 10 |
| [52] | Harvey et al., 2018 | Motor | 199 | Contralesional M1 | 1 | N/A | N/A | 18 |
| [53] | Hsu et al., 2013 | Motor | 12 | Ipsilesional M1 | iTBS (5 Hz bursts/50 Hz pulses) | 1200 | 80 | 10 |
| [54] | Ibrahim et al., 2020 | Motor | 40 | Contralesional dorsal premotor cortex | 5 | 2000 | 80 | 10 |
| [55] | Jorge et al., 2004 | Mood | 20 | Left DLPFC | 10 | 1000 | 110 | 10 |
| [56] | Kim et al., 2014 | Gait | 32 | Ipsilesional cerebellar ataxic side | 1 | 900 | 100 | 5 |
| [57] | Kuzuo et al., 2021 | Motor | 20 | Contralesional M1 | 1 | 1200 | 90 | 10 |
| [58] | Lee et al., 2022 | Speech | 26 | Right pars triangularis | 1 | 1200 | 90 | 5 |
| [59] | Li et al., 2020 | Cognition | 30 | Left DLPFC | 5 | 2000 | 100 | 15 |
| [60] | Li W et al., 2022 | Cognition | 58 | Left DLPFC | iTBS (5 Hz bursts/50 Hz pulses) | 600 | 100 | 10 |
| [61] | Li Y et al., 2022 | Mood | 32 | Left DLPFC | 5 | 2000 | 100 | 20 |
| [62] | Lin et al., 2023 | Speech | 33 | Broca's area | 1 | 900 | 90 | 10 |
| [63] | Liu et al., 2020 | Cognition | 58 | Left DLPFC | 10 | 700 | 90 | 20 |
| [64] | Liu et al., 2024 | Cognition | 100 | Left DLPFC | 10 | 1280 | 90 | 14 |
| [65] | Liu et al., 2022 | Dysphagia | 49 | Ipsilesional mylohyoid cortical region | 5 | 1000 | 80 | 10 |
| [66] | Long et al., 2018 | Motor | 62 | Contralesional M1 | 1 | 1000 | 90 | 15 |
| [67] | Luk et al., 2022 | Motor | 24 | Contralesional M1 | 1 | 1200 | 90 | 10 |
| [68] | Matsuura et al., 2015 | Motor | 20 | Contralesional M1 | 1 | 1200 | 100 | 5 |
| [69] | Meng et al., 2020 | Motor | 28 | Contralesional M1 | 1 | 1200 | 100 | 10 |
| [70] | Park et al., 2013 | Dysphagia | 18 | Contralesional pharyngeal motor cortex | 5 | 500 | 90 | 10 |
| [71] | Park et al., 2017 | Dysphagia | 33 | Mylohyoid muscle area | 10 | 500 | 90 | 10 |
| [72] | Rao et al., 2022 | Dysphagia | 64 | Bilateral cerebellar hemispheres | iTBS (5 Hz bursts/50 Hz pulses) | 600 | 100 | 10 |
| [73] | Seniow et al., 2012 | Motor | 40 | Contralesional M1 | 1 | 1800 | 90 | 15 |
| [74] | Sharma et al., 2020 | Motor | 96 | Contralesional premotor cortex | 1 | 750 | 110 | 10 |
| [75] | Sung et al., 2013 | Motor | 54 | Contralesional M1 | 1 | 600 | 90 | 20 |
| [76] | Tai et al., 2023 | Dysphagia | 90 | suprahyoid muscle cortex | iTBS (5 Hz bursts/50 Hz pulses) | 600 | 80 | 20 |
| [77] | Tang et al., 2024 | Motor | 43 | Ipsilesional sensorimotor network | iTBS (5 Hz bursts/50 Hz pulses) | 600 | 80 | 15 |
| [78] | Tsai et al., 2014 | Speech | 56 | Right pars triangularis | 1 | 600 | 90 | 10 |
| [79] | Wang et al., 2014 | Speech | 45 | Right pars triangularis | 1 | 1200 | 90 | 10 |
| [80] | Wang et al., 2014 | Motor | 48 | Ipsilesional M1 | iTBS (5 Hz bursts/50 Hz pulses) | 600 | 80 | 20 |
| [81] | Wang et al., 2020 | Motor | 44 | Contralesional M1 | 10 | 1000 | 100 | 14 |
| [82] | Wang et al., 2012 | Motor | 24 | Contralesional M1 | 1 | 600 | 90 | 10 |
| [83] | Xie et al., 2021 | Gait | 36 | Contralesional cerebellum | iTBS (5 Hz bursts/50 Hz pulses) | 600 | 80 | 10 |
| [84] | Yang et al., 2021 | Motor | 39 | Ipsilesional M1 | 5 | 750 | 100 | 10 |
| [85] | Yu et al., 2022 | Gait | 18 | Left DLPFC | 5 | 1200 | 80 | 10 |
| [86] | Zheng et al., 2015 | Motor | 112 | Contralesional M1 | 1 | 1800 | 90 | 24 |
| [87] | Zhong et al., 2023 | Dysphagia | 84 | Bilateral pharyngeal motor area | 10 | 250 | 80 | 10 |
| [88] | Zou et al., 2023 | Dysphagia | 45 | Bilateral pharyngeal motor area | 5 | 600 | 90 | 20 |

### Meta analysis by outcome modality

#### Motor

A total of 27 studies assessed participants with motor impairments using the Upper-Extremity Fugl-Meyer Assessment (UEFMA) as the primary outcome measure. Analysis demonstrated that rTMS treatment was overall associated with a significant improvement in motor function compared with sham stimulation (k = 27, MD = 4.68, 95% CI [2.18, 6.54]; z = 4.92, p < 0.0001). Tests of homogeneity indicated significant between-study heterogeneity (Q = 51.4, df = 26, p < 0.005, I² = 49.0%), suggesting that differences in effect sizes across studies exceeded what would be expected from sampling error alone. Moderator analyses were conducted to explore potential sources of heterogeneity.

Subgroup analyses revealed that time since stroke significantly moderated treatment effects (Q = 11.53, df = 2, p < 0.05), suggesting that intervention efficacy varied across acute/subacute, early chronic, and late chronic phases. Post-hoc pairwise contrasts showed that participants in the acute/subacute phase experienced significantly greater improvements in motor function compared with those in the early chronic phase (MD = 5.79, 95% CI [0.54, 11.04], p < 0.05) and the late chronic phase (MD = 6.99, 95% CI [0.76, 13.23], p < 0.05). In contrast, differences between the early chronic and late chronic subgroups were not statistically significant (MD = 1.20, 95% CI [-6.25, 8.66], p = 0.92).

In meta-regression models including stimulation intensity, number of treatment sessions, and participant demographics (age and sex) as continuous moderators, analysis showed no significant moderation of effect size by all moderators: stimulation intensity (z = -1.96, p = 0.051), number of treatment sessions (z = -0.39, p = 0.69), age (z = 0.21, p = 0.83), sex (z = 0.32, p = 0.75). Funnel plot asymmetry tests (z = -1.09, Egger’s p = 0.27) also indicated no evidence of publication bias.

#### Dysphagia

Seven studies assessed participants with dysphagia using the Penetration Aspiration Scale (PAS) as the primary outcome measure. Analysis indicated that rTMS treatment led to a statistically significant improvement in swallowing function compared with sham stimulation (k = 7, MD = -1.50, 95% CI [-2.40, -0.57]; z = -3.18, p < 0.005). The negative mean difference reflects improvement, as lower PAS scores indicate reduced aspiration and better swallowing function. Tests of homogeneity revealed substantial between-study heterogeneity (Q = 131.6, df = 6, p < 0.0001, I² = 92.9%), suggesting that variability in study effect sizes exceeded what would be expected by chance. Subgroup analysis by time since stroke did not reveal significant differences in treatment effects (Q = 2.31, df = 1, p = 0.13). None of the individual moderators featured in meta-regression emerged as significant predictors of effect size (all p > 0.6). Begg’s rank correlation test did not reveal significant funnel plot asymmetry (Kendall’s tau = 0.047, p = 1.00), suggesting no evidence of publication bias in this domain.

#### Language

Five studies assessed participants with speech impairment using the Concise Chinese Aphasia Test (CCAT) as the primary outcome measure. Analysis indicated that rTMS treatment led to a statistically significant improvement in language function compared with sham stimulation (k = 5, MD = 0.62, 95% CI [0.22, 1.01]; z = 3.06, p < 0.005). Tests of homogeneity indicated no significant between-study heterogeneity (Q = 2.89, df = 4, p > 0.05, I² = 0%), suggesting that differences across study effect sizes were consistent with chance variation. Subgroup analysis by time since stroke did not reveal significant differences in treatment effects (Q = 1.47, df = 1, p = 0.22). Begg’s rank correlation test did not reveal significant funnel plot asymmetry (Kendall’s tau = 0.60, p = 0.23), suggesting no evidence of publication bias in this domain.

#### Cognition

Four studies evaluated participants with cognitive impairment using the Mini-Mental State Examination (MMSE). rTMS significantly improved cognitive function compared with sham (k = 4, MD = 1.73, 95% CI [0.84, 2.62]; z = 3.82, p < 0.0001). Tests of heterogeneity showed no meaningful between-study variability (Q = 2.16, df = 3, p > 0.05; I² = 1.0%), indicating that observed differences in effect sizes were largely attributable to chance. Begg’s rank correlation test did not reveal significant funnel plot asymmetry (Kendall’s tau = 0.67, p = 0.33), suggesting no evidence of publication bias in this domain.

#### Gait

Five studies evaluated participants with gait impairments using the 10-Meter Walk Test (10MWT). rTMS did not produce a significant improvement in gait function compared with sham (k = 5, MD = 0.22, 95% CI [-1.27, 1.72]; z = 0.29, p > 0.05). Tests of heterogeneity indicated low between-study variability (Q = 4.85, df = 4, p > 0.05; I² = 34.9%), suggesting that observed differences in effect sizes were modest and consistent with chance variation. Subgroup analysis by time since stroke did not reveal significant differences in treatment effects (Q = 0.0001, df = 1, p = 0.99). Begg’s rank correlation test did not reveal significant funnel plot asymmetry (Kendall’s tau = 0.20, p = 0.82), suggesting no evidence of publication bias in this domain.

#### Post-stroke mood disorders

Four studies evaluated participants with post-stroke depression using the Hamilton Depression Rating Scale 17 (HAMD). rTMS significantly improved mood compared with sham (k = 4, MD = -2.34, 95% CI [-4.38, -0.30]; z = -2.25, p < 0.05). Because lower HAMD scores reflect fewer depressive symptoms, the negative mean difference indicates that participants receiving rTMS experienced greater mood improvement relative to sham. Tests of heterogeneity showed significant between-study variability (Q = 29.2, df = 3, p > 0.05; I² = 88.4%), indicating that observed differences in effect sizes were largely not attributable to chance. Begg’s rank correlation test did not reveal significant funnel plot asymmetry (Kendall’s tau = -0.33, p = 0.75), suggesting no evidence of publication bias in this domain.

**Figure 2.**
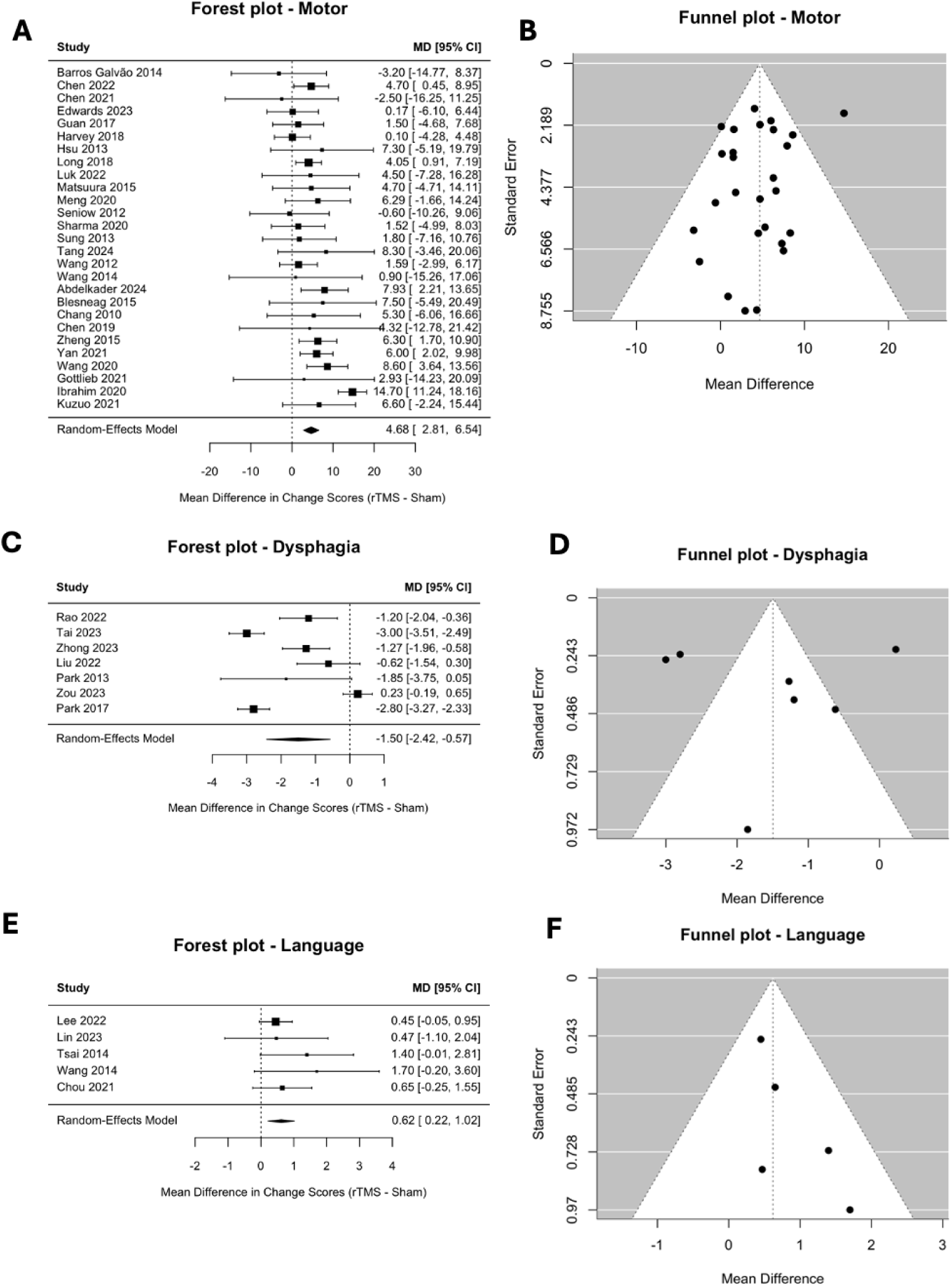
Forrest Plots and Funnel Plots by Outcome Modality. Panel (A) and (B) show forrest and funnel plots for motor impairments, panel (C) and (D) for dysphagia, and panel (E) and (F) for language, respectively.

**Figure 3.**
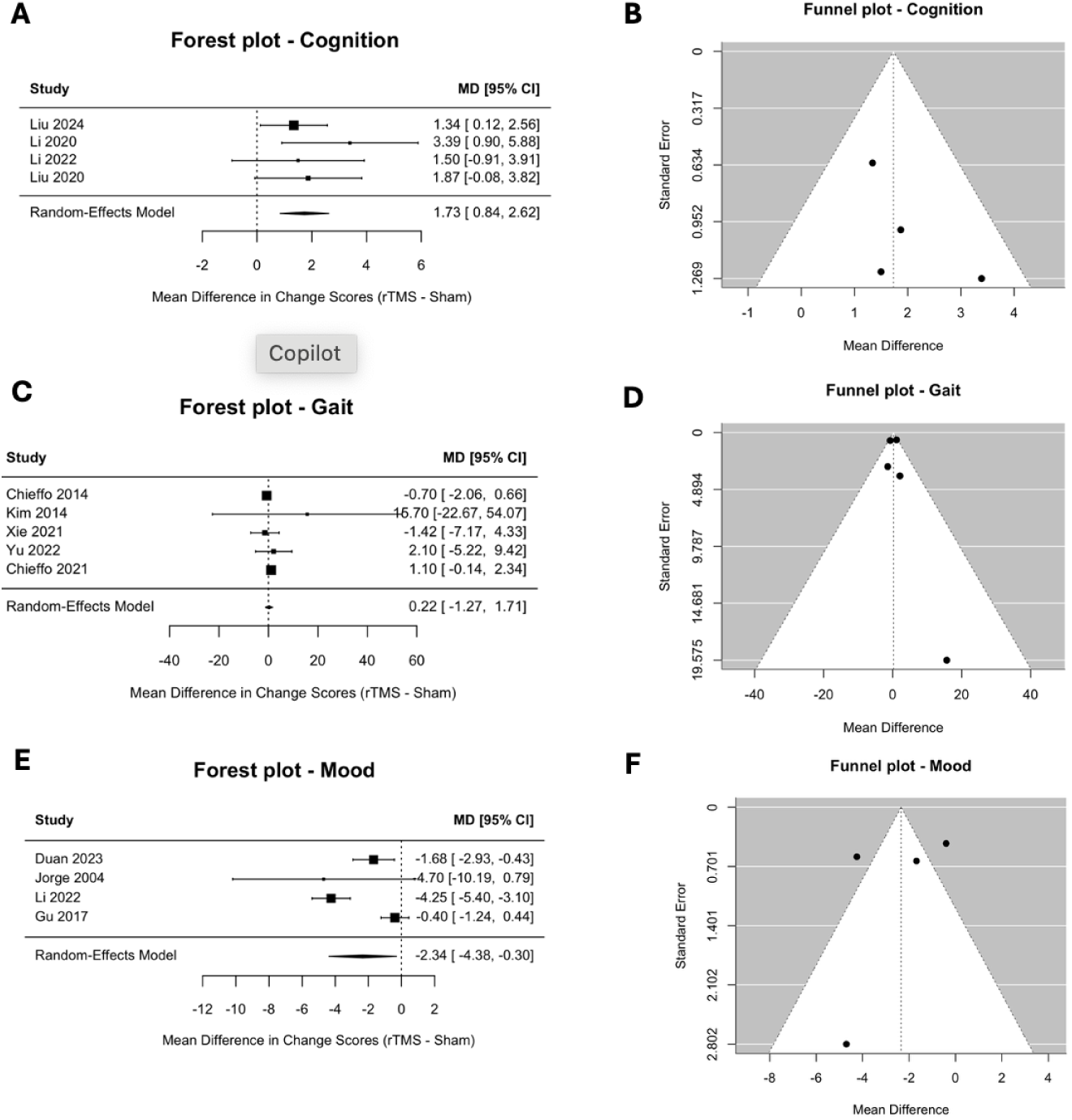
Forrest Plots and Funnel Plots by Outcome Modality. Panel (A) and (B) show forrest and funnel plots for cognition, panel (C) and (D) for gait, and panel (E) and (F) for mood, respectively.

## Discussion

This systematic review and meta-analysis explored the efficacy of rTMS in post-stroke rehabilitation, spanning motor, dysphagia, language, cognition, mood, and gait. Our findings reinforce rTMS as an effective adjunctive intervention, showing significant benefit across all outcome modalities excluding gait. We observed a moderating effect of time since stroke on motor recovery, with RCTs enrolling acute/subacute participants demonstrating significantly greater improvements compared to those including early/late chronic participants. Previous meta-analyses in this area have generally focused on a single outcome domain and have not systematically compared outcomes across different stages of recovery. In contrast, the present meta-analysis expands the literature by synthesizing evidence across multiple functional domains and stratifying effects by time since stroke, allowing for a more comprehensive understanding of both the general efficacy of rTMS in stroke rehabilitation and the potential influence of critical neuroplastic windows on recovery. Our findings align with prior meta-analyses, which consistently demonstrate beneficial effects of rTMS in improving motor function, language abilities, and swallowing, thereby reinforcing the therapeutic potential of rTMS across multiple aspects of post-stroke recovery.

In this review, timing of intervention (acute/subacute, early chronic, late chronic) emerged as a critical determinant in studies on motor recovery. Subgroup analyses demonstrated that the efficacy of rTMS on motor function was significantly moderated by time since stroke (Q = 11.53, df = 2, p < 0.05). Participants in the acute/subacute phase experienced significantly greater gains (MD = 5.79, 95% CI [0.54, 11.04], p < 0.05) compared with those in the early chronic phase, as well as compared with those in the late chronic phase (MD = 6.99, 95% CI [0.76, 13.23], p < 0.05). No differences emerged between early and late chronic participants (MD = 1.20, 95% CI [-6.25, 8.66], p = 0.92). These findings support prior evidence that the early post-stroke period represents a heightened window of neuroplasticity during which neuromodulation may exert stronger therapeutic effects.^33,89^ Animal models and human neuroimaging studies have also demonstrated that cortical excitability and synaptic remodeling are markedly increased during the acute/subacute period following a stroke.^90,91^

Subgroup analyses were conducted for some outcome domains with relatively few studies (e.g., dysphagia, language, gait; k = 5-7). While statistical assumptions for subgroup testing were met, the small number of trials limited statistical power, reducing confidence in the reliability of moderator effects. This reduces the stability of effect size estimates, making the results highly sensitive to the inclusion or exclusion of individual trials. Additionally, tests of heterogeneity are known to be unreliable when applied to outcome modalities with few studies, as they lack the power to detect true between-study variability and may either underestimate or overestimate heterogeneity.^92^ The significant timing effects observed in motor recovery likely reflect the larger pool of available studies in this domain, which provided sufficient power to detect subgroup differences. In all other outcome domains where subgroup analyses were performed, timing of intervention did not significantly moderate treatment effects despite showing significant overall improvements compared to sham stimulation. The absence of timing effects may reflect differences in underlying recovery trajectories. For example, language recovery may follow a more extended or variable timeline than motor recovery, while post-stroke depression may be influenced by psychosocial factors that diminish the relative importance of intervention timing.^93,94^

Previous reviews and meta-analyses have investigated the potential of rTMS in stroke recovery, but these have typically been limited to a single outcome domain, such as motor function, language, or cognition. A majority of reviews and meta-analyses on rTMS in post-stroke rehabilitation focus on motor impairment, specifically upper limb motor recovery. For these studies, meta-analyses have demonstrated rehabilitory effects of rTMS, but none compared effects across timing of administration.^95,96^ Similarly, meta-analyses and reviews of language recovery have shown improvement after rTMS, but have not stratified outcomes by the timing of the intervention.^97,98^ Reviews examining cognitive recovery have summarized effects on global cognition and activities of daily living, typically reporting moderate standardized mean differences with high heterogeneity and limited subgroup analyses.^99^ Reviews examining post-stroke impairments surrounding dysphagia outcomes are limited, often summarizing small sample studies.^100^ Major challenges include heterogeneity in stimulation protocols, smaller sample sizes (often n < 30 per arm), and variable outcome measures, all of which limit generalizability and precision.

Our study is novel in that it is the first to synthesize evidence across all functional domains within a single meta-analysis. Previous analyses are unable to capture the multidimentional nature of stroke recovery, which often involves simultaneous deficits in motor function, language, cognition, swallowing, gait, and mood. This enables systematic comparisons of rTMS efficacy across domains, clarifying whether certain deficits respond more strongly to intervention than others. It also allows for a unified evaluation of moderators, such as time since stroke, and whether they operate similarly across different outcomes. Our study has found that timing of intervention is a significant moderator for motor recovery but not necessarily other outcome domains. This inference is only possible because our synthesis spans multiple domains to detect differences across domains. By aggregating evidence across the full range of post-stroke impairments, our study identifies gaps in the broader applicability of rTMS in post-stroke care. rTMS has been predominantly studied and applied in the context of motor recovery due to its established efficacy in enhancing motor cortex excitability and promoting corticospinal reorganization. However, its potential utility in non-motor domains remains less understood and comparatively underexplored, with existing studies often limited by small sample sizes or heterogeneous protocols.

Findings from this meta-analysis may help explain the variability of results in randomized controlled trials of rTMS in motor recovery. The NICHE trial, a large, multicenter RCT of low-frequency rTMS that enrolled 199 participants 3 to 12 months post-stroke, found no difference between active and sham rTMS when both arms received intensive motor rehabilitation.^52^ Similarly, the E-FIT trial, a large multicenter RCT that used the same 1 Hz rTMS protocol and enrolled 58 participants in the subacute–chronic window, also failed to show a statistically significant advantage of active rTMS versus sham.^48^ Our finding that acute/subacute participants derive larger motor gains than chronic participants provides a plausible explanation for this negative result. Many large RCTs enroll participants in the subacute/chronic window, and if rTMS has its greatest impact early after stroke (when plasticity is heightened), trials dominated by chronic participants may underestimate efficacy.

There are several limitations to this systematic review and meta-analysis. First, many studies were small in size (sample size <40 participants). Second, studies were not balanced for sex or other demographic factors, possibly limiting generalizability of results. Third, there was variability in outcomes evaluated and in methods used to measure them among studies. Fourth, depending on the primary outcome of interest, associated outcomes (i.e. cognitive outcomes in a study of TMS treatment of speech) were secondary and not always reported. Finally, there may be publication bias, which would skew results towards positivity that we are not able to measure. The present findings underscore the need for future rTMS trials to adopt designs that explicitly account for timing of intervention and patient selection. Large-scale RCTs should prioritize early-phase stroke participants and adopt adaptive, multi-arm designs comparing different stimulation parameters. Adaptive trial designs that test multiple stimulation frequencies, intensities, and targets within the same study framework could accelerate discovery of optimal stimulation parameters. By aligning trial design with mechanistic insights and patient-specific predictors, future research can more precisely test the clinical value of rTMS and accelerate its translation into evidence-based rehabilitation practice.

## Conclusions

Our findings support integrating rTMS into acute/subacute stroke rehabilitation protocols, particularly for motor and speech impairments. Timing appears critical: the earlier the intervention, the larger the clinical benefit. Clinicians should consider initiating rTMS-based therapy as soon as patients are medically stable, ideally within the first 12 weeks post-stroke. As protocols become more standardized and individualized dosing algorithms emerge (e.g., via MEP-based predictors or brain imaging), rTMS may transition from a promising modality to a precision therapy in stroke recovery. rTMS holds strong potential as an early-phase enhancer of neuroplastic recovery, especially in motor and speech domains. Realizing its full clinical value will depend on rigorous trials, standardized delivery, and strategic integration with conventional rehabilitation approaches.

## Data Availability

All data referred to in the manuscript is from previously published data that is publicly available.

